# Polygenic risk scores for autoimmune related diseases are significantly different and skewed in cancer exceptional responders

**DOI:** 10.1101/2023.02.22.23285773

**Authors:** Siyuan Chen, Amelia LM Tan, Cassandra Perry, Susanne Churchill, Margaret Vella, Jenny Mao, Vinay Viswanadham, Shilpa Kobren, Isaac S Kohane

## Abstract

A group of 54 exceptional responders (ERs) to cancer treatment across a variety of cancers and treatments were compared to typical cancer patients using previously defined polygenic risk scores (PRS) for multiple autoimmune-related diseases including type 1 diabetes (T1D), hypothyroidism, psoriasis, rheumatoid arthritis, multiple sclerosis, and inflammatory bowel (IBD) disease. Most of the ERs were not treated with checkpoint inhibitors and included a broad array of tumor types. Significantly elevated PRSs were found between ERs relative to typical cancer patients in T1D, hypothyroidism, and psoriasis. IBD PRS scores were significantly decreased in the ERs.

## Introduction

Even in cancer trials that have failed to meet their pre-specified endpoints there sometimes are individuals who have outcomes that exceed the most optimistic expectations, distinguishing these patients as remarkable outliers. In more successful trials, there are patients who exceed the mean response of the intervention arm thus categorizing them as outliers. Historically, these outliers have not been paid much attention, but with the advent of whole exome or whole genome sequencing (WES, WGS), interest in mechanisms responsible for the remarkable outcomes—exceptional responders (ER)—has grown considerably. By definition, the study cohorts are composed of rare patients and are therefore small (in the low hundreds of participants[1]) or sometimes consist of single case reports[2–4]. The rarity of these patients has encouraged researchers to analyze highly heterogeneous groups, covering a variety of cancers and treatments[1,5–7]. Important insights, mostly from somatic cancer genomes and less often from germline genomes, have been obtained in these studies about mechanisms that might underlie exceptional responses in cancer, and they have been as heterogeneous as the patients. These mechanisms include DNA repair, signaling, and the endogenous variations in immune response[8]. For example, a *BRCA1* germline gene deletion was found in a patient with recurrent ovarian cancer, who showed exceptional response to Olaparib[2]. In another case, two patients with breast cancer and different germline *PTEN* mutations both showed an exceptional response to the AKT inhibitor, capivasertib[3]. The mechanistic studies included in these reports are compelling but they only account for a small fraction of the ERs.

In 2018, we launched the registry for the Network for Enigmatic Exceptional Responders (NEER) across all trials as a patient directed study so that any patient could volunteer their own case and data as a candidate for the NEER registry. The first 56 patients who met the criteria (see methods) were enrolled and sequenced. Consequently, the registry contains a broad collection of cancers (see Table 1) as well as a range of therapies from chemotherapy, radiation therapy and immunotherapy (see Supplementary Table 1). Here we describe the results of our first study across the germline genomes of NEER participants, looking for common genomic predispositions to exceptional response. Since the contribution to an exceptional response of any given common variant is going to be small, we re-used previously published polygenic risk scores (PRS) to evaluate whether ERs have PRSs that are significantly different from those of typical cancer patients. Prior studies have reported that elevated PRSs for autoimmune diseases such as hypothyroidism in patients receiving immunotherapy are associated with autoimmune adverse events and variably associated with improved outcomes [9,10]. Rather than generating a new set of scores based on a single reference population, we decided to use previously published PRSs to be able to directly compare findings in NEER with the claims of the prior studies. The hypothesis articulated in those reports was that autoimmunity led to better outcomes by modulating overall reactivity of helper T cells and B cells. Therefore, we broadened the PRSs studied to 6 autoimmune-related diseases and multiple cancer therapies to determine what kinds of autoimmune or inflammatory processes were associated with improved outcomes and whether the improvement was present in chemotherapies and other treatments not classically defined as immunomodulating.

**Table 1.** Demographic summary of NEER and PCAWG subset.

| Characteristics | NEER (n=54) | PCAWG (n=1376) |
| --- | --- | --- |
| Age |  |  |
| Mean (range) | 66.1 (39-87) | 54.7 (1-90) |
| Median (range) | 66 (39-87) | 59 (1-90) |
| Female sex — no.(%) | 33 (61.1%) | 634 (46.1%) |
| Cancer type |  |  |
| Breast — no.(%) | 13 (24.1%) | 123 (8.9%) |
| Pancreatic — no.(%) | 8 (14.8%) | 245 (17.8%) |
| Lung — no.(%) | 6 (11.1%) | 37 (2.7%) |
| Others — no.(%) | 27 (50.0%) | 971 (70.1%) |
| Multiple cancer — no.(%) | 10 (18.5%) | - |
| Cancer stage |  |  |
| Stage 1 — no.(%) | 0 (0%) | 296 (21.5%) |
| Stage 2 — no.(%) | 3 (5.6%) | 82 (6.0%) |
| Stage 3 — no.(%) | 2 (3.7%) | 228 (16.6%) |
| Stage 4 — no.(%) | 27 (50.0%) | 90 (6.5%) |
| NAs — no.(%) | 21 (38.9%) | 680 (49.4%) |
| Survival time (censored) |  |  |
| Mean (range) | 5086 (515-12477) | 1311 (0-10958) |
| Median (range) | 4083 (515-12477) | 834 (515-12477) |

## Methods

### Cancer exceptional responders sample collection

NEER exceptional responders were obtained from a group of US-based applicants aged at least 18 years old with an exceptional response to cancer therapies. An exceptional response was generally considered to be 2 standard deviations greater than the most recently available survival means, estimated separately per cancer type. Of 222 individuals who registered for the study, 82 were accepted based on their eligibility. Eligibility was determined based on the most recently available survival means within each cancer type, with eligible participants exceeding 2 standard deviations greater than the survival rate or exhibited significant deviation from standard clinical treatment. The participants were further evaluated by cancer-specific oncology experts (see Acknowledgements) to ensure that the ER candidates were indeed outliers, based on their presentation and course, before they were enrolled. Electronic health records and blood samples were collected from 56 ERs at the time of this analysis. Whole blood samples of participants were used for whole genome sequencing (WGS) with 30X mean coverage at the Broad Institute. DNA was extracted from aliquots of whole blood using the QIAsymphony DSP DNA Kit in conjunction with the QIAsymphony SP instrument (Qiagen). DNA was processed for PCR-Free library construction, sequenced with 150bp paired-end reads, and sample identification QC check. A KAPA HyperPrep library preparation was followed by qPCR quantification.

### Data processing

WGS data was processed through the CGAP pipeline developed by Harvard’s Department of Biomedical Informatics and Brigham Genomic Medicine. Variants were filtered through GQ > 20 and VQSR as quality controls. The NEER dataset assembly is based on the GRCh38 genome but most PRS models were from GRCh37. A liftover procedure [11] was used to convert datasets from one genome assembly to another to make them comparable. Minimac4 1.5.7 genotype imputation[12] was applied, based on Haplotype Reference Consortium (HRC) panel, to increase power and improve the PRS [13,14]. An ancestry population check was performed through PCA based on all shared variants between NEER ERs and PCAWG typical cancer patients after NEER data went through the genomic liftover procedure and was imputed.

Two NEER participants (55 and 56) were found to be distant from a cluster of the other 54 participants in principal component analyses based on SNPs (Supplementary Figure 1). They also carried the largest number of variants and the largest number of homozygous variants and were identified as having a non-European continent of origin. Earlier PRS models have been shown to be brittle in their performance when applied to populations differing from those they were developed on[15–19] even though recent methods and data sources are resulting in greater robustness[20,21]. As most of the PRS models were developed on European populations, only the 54 NEER ERs were included.

The germline jointly genotyped WGS dataset of PCAWG went through the same quality control filter and only the population of European origin was retained. Since patients in PCAWG annotated as having a complete remission, partial remission or no evidence of disease may share some similar characteristics with ERs, they were removed from the comparison group. After filtering, we obtained a 1376-patient pan-cancer control dataset.

### Polygenic risk score calculation

PRS is calculated as a sum of weighted effect alleles. The general mathematical formula of the PRS is written as follows:

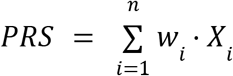

where X_i_ denotes the effect allele count and w_i_ denotes the weight of the i-th SNP for a specified outcome. The number of SNPs included in PRS varies, depending on the trait/disease and were determined in the earlier studies[9,22,23] (see Supplementary Table 2). When comparing PRSs between NEER and PCAWG, only variants shared between the two cohorts were included. The PRS models implemented and the number of SNPs in each model are listed in the Supplementary Table 2.

### Survival analysis

A Kaplan-Meier (KM) analysis was performed among the selected PCAWG patients. Patients with missing data in vital status were excluded. Patients were divided into high risk groups and low risk groups based on the 10% and 90% quantile of PRSs.

### Statistical analysis

A log rank test was used to compare the difference in survival time in the KM curve. Wald tests of logistic regression coefficients were used as statistical tests between two groups of PRSs. All tests were two-tailed tests at α < 0.05 level.

### Gene set enrichment analysis

SNPs in each PRS model were annotated with credible regions based on LD-annot [24]and the genes located in the regions were collected. Genes were then ranked by gene level score: ∑ β_*i*_ N_*i*_ where N_*i*_ is the number of alleles in the NEER cohort for each SNP and β_*i*_ is the coefficient for the corresponding SNP in the PRS., The R package fgsea[25] was used to compute gene set enrichment analysis based on the GO Biological Process (GOBP) ontology.

### Skew Analysis

While the difference in distribution test above compares PCAWG to NEER, it does not test whether the PRS scores are skewed within each population individually. The estimate of skew is based on the sample skewness[26] b_1_ where

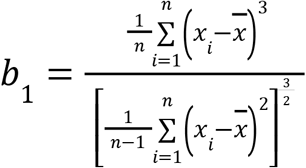

This estimate was calculated for PCAWG and NEER using the skewness.norm.test function in the normtest package in R.

## Results

### Characteristics of the NEER Exceptional Responders

The NEER participants were 56 cancer ERs with different cancer types. Of these 56 participants, a majority of them were breast cancer patients (Table 1). ERs underwent a variety of therapies, including antiangiogenic therapy, chemotherapy, hormone therapy, immunotherapy, radiation and targeted therapy. A heterogeneous set of chemotherapies protocols were administered to 73.2% of the patients. Of the latter, antimetabolite drugs and anti-mitotic drugs were administered to 18 ERs (Table 1). Other than radiation, paclitaxel was the chemotherapeutic drug assigned to the most patients, followed by gemcitabine and carboplatin, which were assigned to 13, 11 and 9 ERs respectively (Supplementary Table 1). Immunotherapy including checkpoint inhibitors was administered to 7 patients (12.5%)

The two outliers in NEER ERs were excluded in the following analysis and a comparable PCAWG control set was identified according to population and response to treatments (see methods). PCAWG was selected as it comprises a large heterogeneous set pan-cancer cohort with Whole Genome Sequencing. For comparison, ERs in NEER were older and had higher (more advanced) cancer stages compared to typical cancer patients, but they lived about 4-times longer.

### Differences in PRSs between NEER and PCAWG

We implemented several previously published PRSs models corresponding to several autoimmune/inflammatory disorders, including type 1 diabetes (T1D)[22], rheumatoid arthritis (RA)[18], psoriasis[18], multiple sclerosis (MS)[23], inflammatory bowel disease (IBD)[19] and hypothyroidism[9]. The ERs showed significantly higher PRSs for hypothyroidism, T1D and Psoriasis risk. In contrast there were significantly lower PRSs for IBD risk compared to typical cancer patients (Figure.1). RA and MS PRSs were not significantly different.

These significant differences in PRS were not reflected in the clinical history of the ERs who had no record of autoimmune disease or inflammatory disease. For each of the PRSs we also checked the survival curves of PCAWG patients when stratified by high vs low PRS (see Methods). These are illustrated in Supplementary Figure 2. None of the differences were statistically significant.

**Figure 1.**
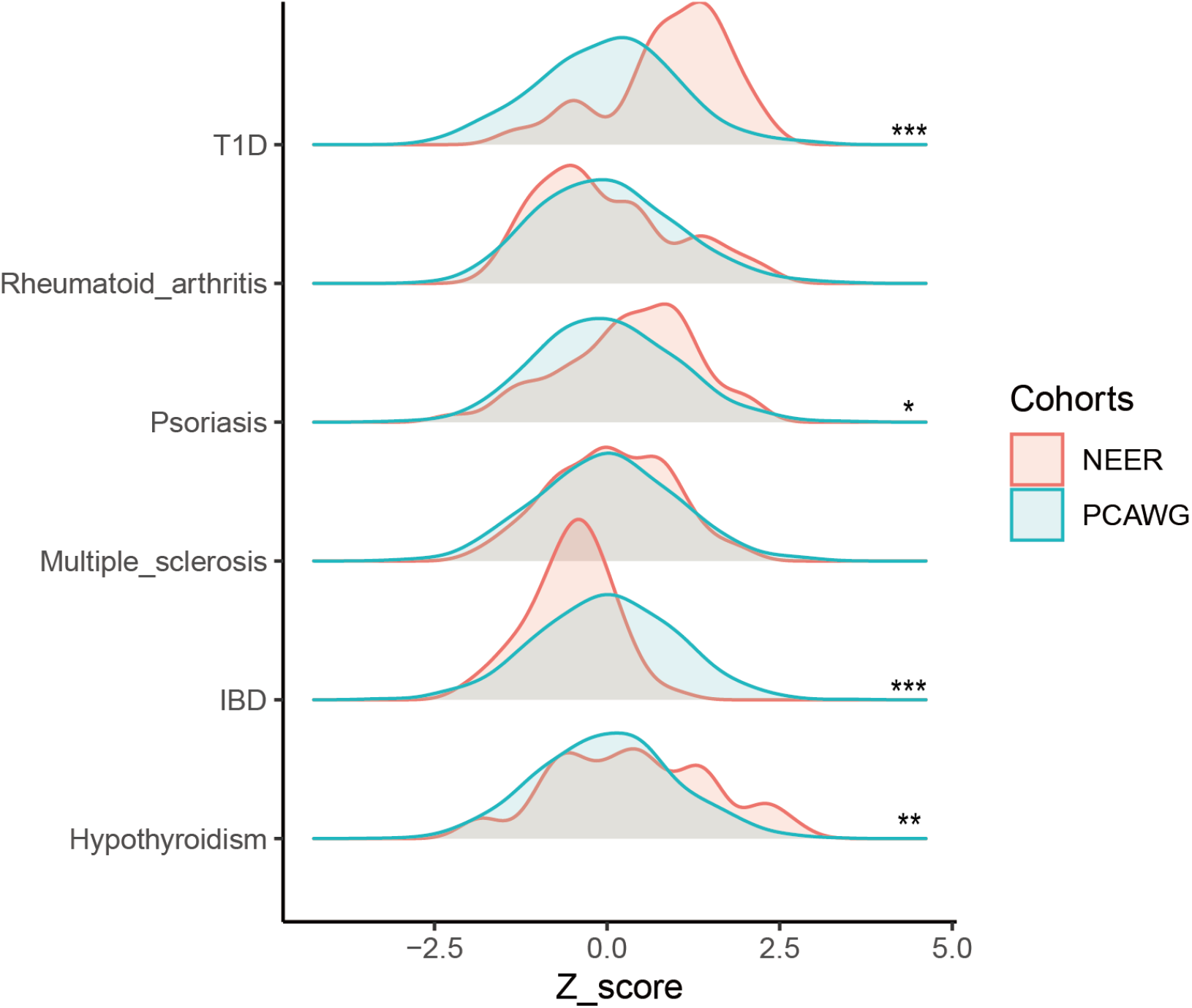
Distribution of PRS Scores in NEER and PCAWG across 6 autoimmune/inflammatory-related disorders. The asterisks denote significance per the Wald test. * signifies p<0.05 ** p < 0.01 *** p < 0.001

The tests in PRS distribution illustrated in Figure 1 compare the PCAWG cohort to NEER. We also determined if the scores within each cohort were significantly skewed from the normal distribution. Despite the much smaller size of the NEER cohort, when we tested for skew in the distribution of PRSs, T1D score skew reached significance (see Table 2) with p = 0.008 whereas PCAWG did not (*p* = 0.433). Skew for all the other NEER PRSs was not significant. PCAWG patients had a highly significant right-skew for both RA and psoriasis (p < 2.2 e-16). The NEER T1D PRS distribution was left-skewed, which indicates that many NEER patients were at higher risk of T1D than a typical cancer patient.

**Table 2.**
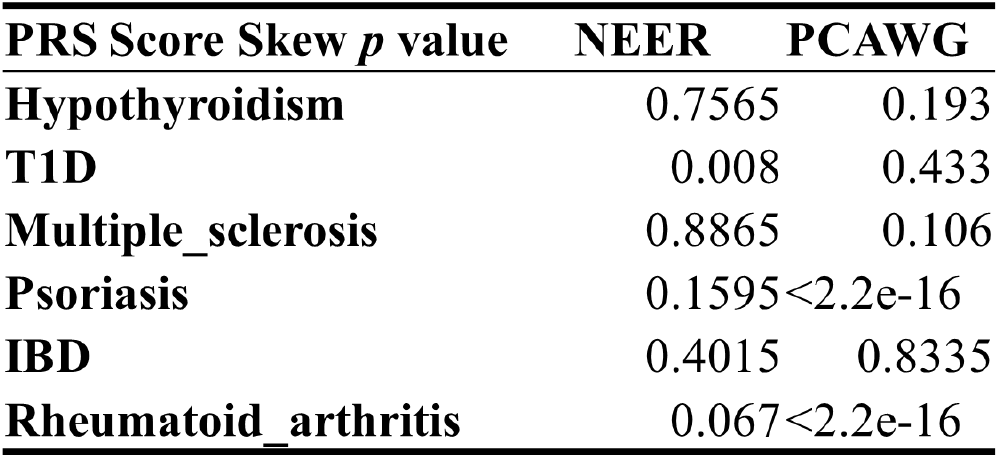
Skew of distribution from normal in PRS of each population.

| **PRS Score Skew $p$ value** | **NEER** | **PCAWG** |
| --- | --- | --- |
| **Hypothyroidism** | 0.7565 | 0.193 |
| **T1D** | 0.008 | 0.433 |
| **Multiple_sclerosis** | 0.8865 | 0.106 |
| **Psoriasis** | 0.1595 | $<2.2 \times 10^{-16}$ |
| **IBD** | 0.4015 | 0.8335 |
| **Rheumatoid_arthritis** | 0.067 | $<2.2 \times 10^{-16}$ |

The top ranked enriched pathways of the four significant PRS autoimmune disorders were calculated by gene set enrichment analysis[25]. Only one of the PRS had statistically significant enriched pathways, IBD, with a p = 0.015 after multiple hypothesis correction for a large number of pathways tested. The enriched pathways for the IBD PRS implicated cell morphology, cell migration and development (e.g. genes *DAB1, ROBO1*, see Supplementary Table 3 for the GSEA ranked pathways). Hypothyroidism, and psoriasis had top ranked pathways in immune signaling (e.g. *PTPN22, IL23R, IL27*) and immune cell activation and cytotoxicity (e.g. *TYK2, IL12B)* but none reached significance. T1D’s leading enriched pathways included metabolic signaling genes like insulin and tyrosine hydroxylase but also did not reach significance after multiple hypothesis testing.

## Discussion

We collected germline WGS data from 56 ERs with different cancer types and treatments and analyzed the combined effect of multiple germline common variants in ERs. Specifically, previously developed PRS scores for autoimmune or inflammatory disease NEER participants were compared to those of a more typical cancer cohort: PCAWG. Two NEER patients were removed from the analysis as their common variant profiles diverged from those of European origin, for which most published PRS profiles have been computed, although this is likely to change soon[20,21]. Within the remaining 54 ERs, there was a significant enrichment of elevated PRS for hypothyroidism and psoriasis based on PRSs in ERs as compared to typical cancer patients. This is consistent with previous studies of the response to immune checkpoint inhibitors and cancer overall survival as well as autoimmune adverse events[9,27] and extends it to patients who have only received chemotherapy or radiotherapy.

Significantly increased T1D PRSs and decreased IBD PRSs were found in ERs, which suggests different immune responses between ERs and typical cancer patients. The divergence in risk profiles for the same common variants between IBD and other autoimmune diseases like T1D has been previously noted and summarized in a study by Wang [28] at multiple loci. The association in opposite directions was observed in the protein tyrosine phosphatase non-receptor type 22 gene (*PTPN22*) which appeared in several of the leading edge gene lists from the gene set enrichment tabulated in Supplementary table 3 as well as multiple independent MHC loci. In those studies divergence was with respect to autoimmune disease, not cancer outcome. Whether the divergence arises from a different immune set point or different (micro)environmental stimuli remains unknown.

There are 66 SNPs in the T1D model and 4 of them were nonsynonymous exonic variants. The genes involved are SH2B3, APOBR, TYK2 and SIRPG. TYK2 is associated with heterodimeric/multimeric cytokine receptors, and thus is involved in cancer immunity[29]. However, when comparing the allele frequencies of the 66 SNPs between NEER and PCAWG, no significant difference was found. Taken together with the skewed T1D PRS distribution, it suggests an accumulation effect of these common variants contributes to the difference rather than the effects of a single gene.

Prior studies of the impact of elevated or decreased PRS scores have focused on checkpoint inhibitors, specifically PD-1/PD-L1 blockage. The results have consistently shown association with new onset autoimmune adverse events (e.g. hypothyroidism, colitis, hypophysitis) but have been less consistent in relating these to outcomes. A study of non-small cell lung cancer by Luo et al. [10] did not reveal that a high PRS for hypothyroidism resulted in a benefit to survival. In contrast, in a study by Khan et al.[9] there was a survival benefit for patients with triple negative breast cancer. Another study by Khan et al. [27] showed long overall survival for patients with bladder cancer high PRS for psoriasis and low PRS for atopic dermatitis.

Unlike the aforementioned studies, the majority of NEER patients (93%) were not treated with checkpoint inhibitors. Yet, over the NEER patients’ wide range of treatments and cancers there were still significant differences in the distribution of PRSs for hypothyroidism and psoriasis. In addition, the ERs had significantly higher T1D PRSs and significantly lower scores for IBD than PCAWG typical cancer patients. Comparing survival times for high and low PRS scores in these same autoimmune/inflammatory diseases did not reveal any significant difference in survival.

By design this study focused on a hypothesis, articulated by prior studies, about germline differences in common variants in autoimmune disease risk. It therefore cannot account for differences in the somatic genome of the tumor, nor those of rare variants which individually might have greater effect sizes for individuals. It also does not address differences in lifestyle and socioeconomic status which is documented for NEER patients but not the large comparison populations like PCAWG. Nonetheless, the reproducibility of the PRS findings from earlier studies and significant PRS findings for additional autoimmune diseases such as T1D in a dataset containing various tumor types and cancer therapies support the hypothesis of a mechanistic link from an altered set point or specificity of immune surveillance and cancer response to treatment in exceptional responders.

## Data Availability

All SNP data and diagnoses produced in the present study are available upon request to the authors.

## Acknowledgements

We are grateful to the following expert oncologists who helped us evaluate candidate ERs: Dr. David Fisher, Dr. Alice Shaw, Dr. Mark Johnson, and Dr. Funda Meric-Bernstam. We are also thankful for Professors Shamil Sunyaev and Professors Peter Park’s comments which were helpful in improving the manuscript.

## Supplementary Figure

**Supplementary Figure 1.**
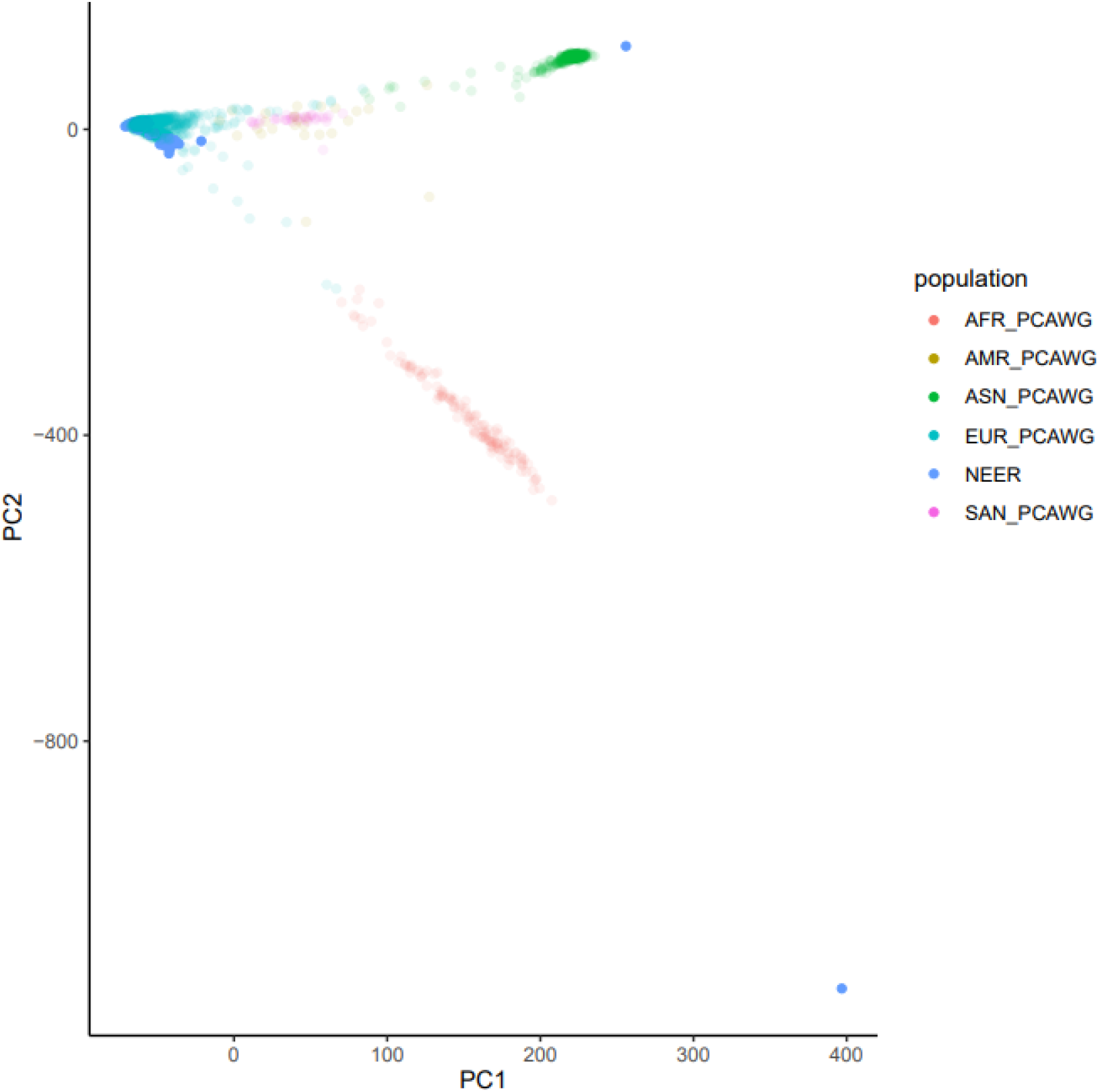
2-D PCA plot of all NEER and PCAWG patients.

**Supplementary Figure 2.**
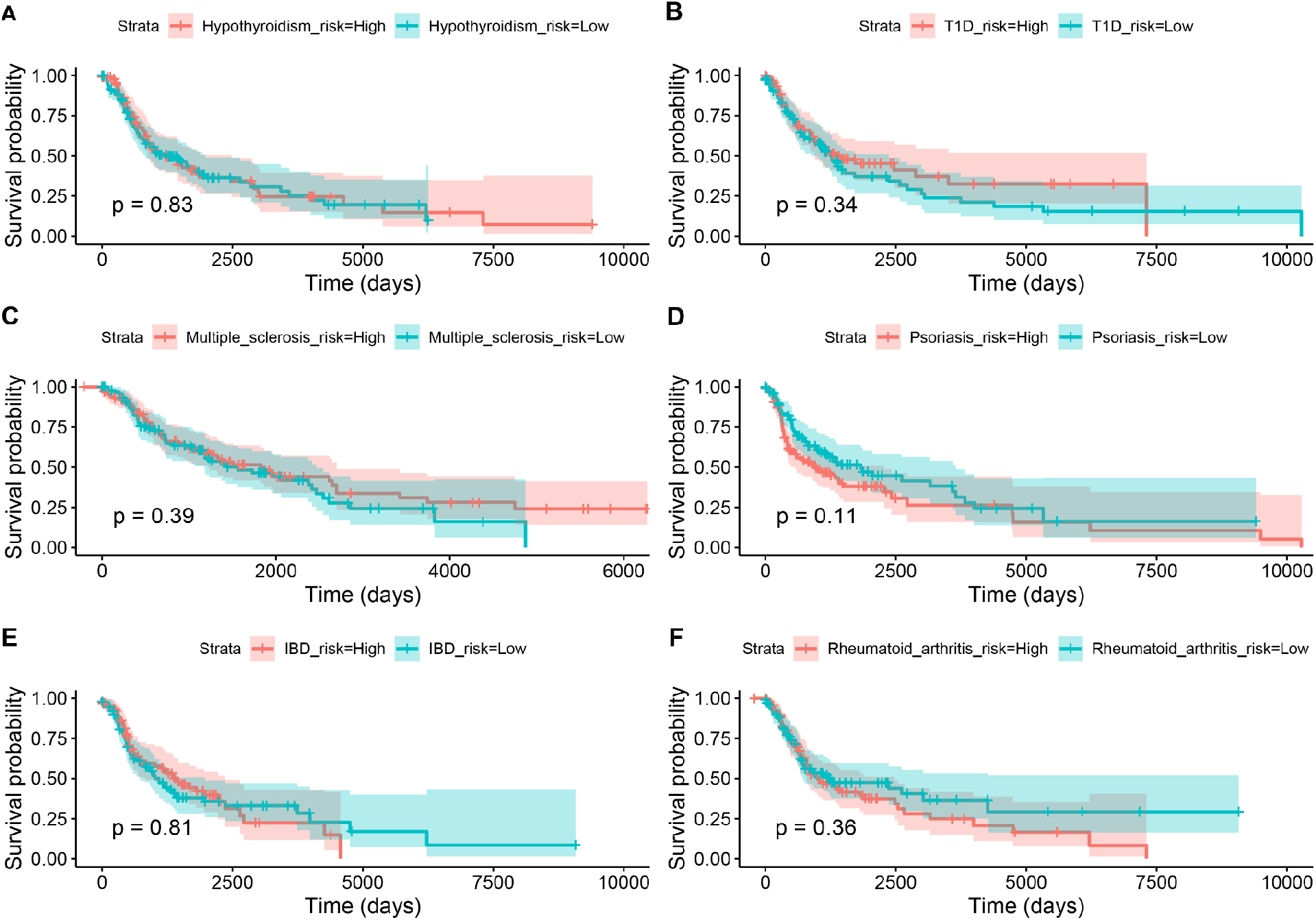
Survival curves of PCAWG patients stratified by high vs low PRS score.

